# Estimation of incubation period distribution of COVID-19 using disease onset forward time: a novel cross-sectional and forward follow-up study

**DOI:** 10.1101/2020.03.06.20032417

**Authors:** Jing Qin, Chong You, Qiushi Lin, Taojun Hu, Shicheng Yu, Xiao-Hua Zhou

## Abstract

**Background:** The current outbreak of coronavirus disease 2019 (COVID-19) has quickly spread across countries and become a global crisis. However, one of the most important clinical characteristics in epidemiology, the distribution of the incubation period, remains unclear. Different estimates of the incubation period of COVID-19 were reported in recent published studies, but all have their own limitations. In this study, we propose a novel low-cost and accurate method to estimate the incubation distribution.

**Methods:** We have conducted a cross-sectional and forward follow-up study by identifying those asymptomatic individuals at their time of departure from Wuhan and then following them until their symptoms developed. The renewal process is hence adopted by considering the incubation period as a renewal and the duration between departure and symptom onset as a forward recurrence time. Under mild assumptions, the observations of selected forward times can be used to consistently estimate the parameters in the distribution of the incubation period. Such a method enhances the accuracy of estimation by reducing recall bias and utilizing the abundant and readily available forward time data.

**Findings:** The estimated distribution of forward time fits the observations in the collected data well. The estimated median of incubation period is 8·13 days (95% confidence interval [CI]: 7·37-8·91), the mean is 8·62 days (95% CI: 8·02-9·28), the 90th percentile is 14·65 days (95% CI: 14·00-15·26), and the 99th percentile is 20·59 days (95% CI: 19·47, 21·62). Compared with results in other studies, the incubation period estimated in this study is longer.

**Interpretation:** Based on the estimated incubation distribution in this study, about 10% of patients with COVID-19 would not develop symptoms until 14 days after infection. Further study of the incubation distribution is warranted to directly estimate the proportion with long incubation periods.

**Funding:** This research is supported by National Natural Science Foundation of China grant 8204100362 and Zhejiang University special scientific research fund for COVID-19 prevention and control.

**Research in context:** *Evidence before this study:* Before the current outbreak of coronavirus disease (COVID-19) in China, there were two other coronaviruses that have caused major global epidemics over the last two decades. Severe acute respiratory syndrome (SARS) spread to 37 countries and caused 8424 cases and 919 deaths in 2002-03, while Middle East respiratory syndrome (MERS) spread to 27 countries, causing 2494 cases and 858 deaths worldwide to date. Precise knowledge of the incubation period is crucial for the prevention and control of these diseases. We have searched PubMed and preprint archives for articles published as of February 22, 2020, which contain information about these diseases by using the key words of “COVID-19”, “SARS”, “MERS”, “2019-nCoV”, “coronavirus”, and “incubation”. We have found 15 studies that estimated the distribution of the incubation period. There are four articles focused on COVID-19, five on MERS, and six on SARS. Most of these studies had limited sample sizes and were potentially influenced by recall bias. The estimates for mean, median, and percentiles of the incubation period from these articles are summarized in Table 1.

*Added value of this study:* In the absence of complete and robust contact-tracing data, we have inferred the distribution of the incubation period of COVID-19 from the durations between departure from Wuhan and symptom onset for the confirmed cases. More than 1000 cases were collected from publicly available data. The proposed approach has a solid theoretical foundation and enhances the accuracy of estimation by reducing recall bias and utilizing a large pool of samples.

*Implications of all the available evidence:* Based on our model, about 10% of patients with COVID-19 do not develop symptoms until 14 days after infection. Further study of individuals with long incubation periods is warranted.

## Introduction

The Center for Disease Control and Prevention (CDC) of China and World Health Organization (WHO) are closely monitoring the current outbreak of coronavirus disease 2019 (COVID-19). It was first identified in Wuhan, Hubei province, China, and has quickly spread across countries and become a global crisis. As of February 22, 2020, the National Health Commission (NHC) of China had confirmed a total of 76 936 cases of COVID-19 in mainland China, including 2442 fatalities and 22 888 recoveries.^1^ Various containment measures, including travel restrictions, isolation, and quarantine have been implemented in China with the aim to minimize virus transmission via human-to-human contact.^2^

**Table 1.** Estimates for the incubation periods of SARS, MERS, and COVID-19.

| Incubation distribution metric | SARS | MERS | COVID-19 |
| --- | --- | --- | --- |
| Mean (SD) or Mean (95% CI) | Hong Kong:<sup>7</sup> 4.4 (4.6);<br>Beijing:<sup>7</sup> 5.7 (9.7);<br>Taiwan:<sup>7</sup> 6.9 (6.1);<br>Hong Kong:<sup>8</sup> 4.6 (15.9);<br>Mainland China:<sup>9</sup> 5.29 (12.33);<br>Singapore:<sup>10</sup> 4.83 (4.37-5.29);<br>Hong Kong:<sup>11</sup> 6.37 (5.29-7.75) | Saudi Arabia:<sup>13</sup> 5.0 (4.0-6.6);<br>South Korea:<sup>13</sup> 6.9 (6.3-7.5);<br>South Korea:<sup>14</sup> 6.9 (6.3-7.5);<br>Saudi Arabia:<sup>14</sup> 5.0 (4.4-6.6);<br>South Korea:<sup>14</sup> 6.7 (6.1-7.3) | Wuhan:<sup>3</sup> 5.2 (4.1-7.0);<br>Mainland China:<sup>5</sup> 5.8 (4.6-7.9);<br>Global:<sup>6</sup> 5.6 (5.0-6.3) |
| Median or Median (95% CI) | Hong Kong, Canada, & USA:<sup>12</sup> 4 | South Korea:<sup>15</sup> 6.0 (4.0-7.0);<br>Middle East:<sup>16</sup> 5.2 (1.9-14.7);<br>South Korea:<sup>17</sup> 6.3 (5.7-6.8) | Mainland China:<sup>4</sup> 3.0 |
| Percentiles | Mainland China, 90%:<sup>9</sup> 10.7;<br>Hong Kong, Canada, & USA, 90%:<sup>12</sup> 12;<br>Singapore, 95%:<sup>10</sup> 9.66 (0.5);<br>Mainland China, 95%:<sup>9</sup> 13.91;<br>Hong Kong, 95%:<sup>11</sup> 14.22;<br>Mainland China, 99%:<sup>9</sup> 20.08 | NA | Mainland China, 2.5%:<sup>5</sup> 1.3;<br>Wuhan, 95%:<sup>3</sup> 12.5;<br>Mainland China, 97.5%:<sup>5</sup> 11.3 |

COVID-19=coronavirus disease 2019. MERS=Middle East respiratory syndrome. SARS=severe acute respiratory syndrome.

Quarantine of individuals with exposure to infectious pathogens has always been an effective approach for containing contagious diseases in the past. One of the critical factors to determine the optimal quarantine of asymptomatic individuals is a good understanding of the incubation period, and this has been lacking for COVID-19.

The incubation period of an infectious disease is the time elapsed between infection and appearance of the first symptoms and signs. Precise knowledge of the incubation period would help to provide an optimal length of quarantine period for disease control purpose, and also is essential in the investigation of the mechanism of transmission and development of treatment. For example, the distribution of the incubation period is used to estimate the reproductive number *R*, that is, the average number of secondary infections produced by a primary case. The reproductive number is a key quantity that impacts the potential size of an epidemic. Despite the importance of the incubation period, it is often poorly estimated based on limited data.

To the best of our knowledge, there is only a handful of studies estimating the incubation period of COVID-19. Among them are Li et al (2020), Guan et al (2020), Backer et al (2020), and Linton et al (2020).^3–6^ In Li et al, the first 425 lab-confirmed cases, reported as of January 22, 2020, were included in the study, while only ten cases could be identified with the exact dates of exposure.^3^ The distribution of the incubation period was subsequently approximated by fitting a lognormal distribution to these ten data points, resulting in a mean incubation period of 5·2 days (95% CI: 4·1-7·0), and the 95th percentile is 12·5 days. However, given the limited sample size, it is challenging to make a solid inference on the distribution of the incubation period. A different result was reported by Guan et al, based on 291 patients who had clear information regarding the specific date of exposure as of January 29, 2020, stating that the median incubation period was 4·0 days (interquartile range, 2 to 7).^4^ However, such study of the incubation period can be highly influenced by the individuals’ recall bias or interviewers’ judgement on the possible dates of exposure rather than the actual dates of exposure that, in turn, might not be accurately monitored and determined, thus leading to a high percentage of error. In Backer et al, 88 confirmed cases detected outside Wuhan were used to estimate the distribution of the incubation period.^5^ For each selected case, a censored interval for the incubation period can be obtained by travel history and symptoms onset. The distribution of the incubation period can then be estimated by fitting a Weibull, gamma, or lognormal distribution with censored data. However, this method contained two types of sampling biases: (1) with the longer incubation period, the patients who resided at Wuhan but developed symptoms outside Wuhan were easier to be observed and hence lead to an overestimation; (2) if the follow-up time (from infection to the end of the study) is short, only the shorter incubation period would be observed and hence lead to an underestimation. Linton et al proposed a similar approach to estimate the incubation period of Backer et al, but corrected the aforementioned second sampling bias.^5,6^ However, the first problem in regard to sampling bias is still an unsolved issue. The estimates of the incubation period from these four studies, together with other results of two other coronavirus disease, SARS and MERS, are listed in Table 1.

To overcome the aforementioned problems, we propose a novel method to estimate the incubation period of COVID-19 by using the well-known renewal theory in probability.^18^ Such a method enhances the accuracy of estimation by reducing recall bias and utilizing abundance of the readily available forward time with a large sample size of 1211. To the best of our knowledge, this paper is a study of the distribution of the incubation period involving the largest number of samples to date. We find the estimated median of the incubation period is 8·13 days (95% CI: 7·37-8·91), and mean is 8·62 days (95% CI: 8·02-9·28), the 90th percentile is 14·65 days (95% CI: 14·00-15·26), and the 99th percentile is 20·59 days (95% CI: 19·47-21·62). Our estimated incubation period of COVID-19 is longer than the those given by previous researches on SARS, MERS, and COVID-19 in Table 1.

## Methods

### Motivations

As described in the previous section, the distribution of the incubation period in most of the literature is either described through a parametric model or its empirical distribution based on the observed incubation period from the contact-tracing data. However, the contact-tracing data are challenging and expensive to obtain, and their accuracy can be highly influenced by recall bias. Hence, a low-cost and high-accuracy method to estimate the incubation distribution is needed.

Based on the COVID-19 daily updates from provincial and municipal health commissions in China, we notice that there is an abundance of cases who asymptomatically left Wuhan, the epicenter of COVID-19, and developed symptoms outside Wuhan. Assuming that these cases were infected before their departure from Wuhan, the time differences between departure and symptoms onset is the censored observations of their incubation periods. Hence, we conducted a cross-sectional and forward follow-up study by assuming to catch those asymptomatic individuals at their departure time and followed them until their symptoms developed. Using the language of renewal processes, we can treat the development of the disease starting from infection by a pathogen as a stochastic process that could be observed from a specific time point in chronological order. In this study, the specific time point refers to the time of departure from Wuhan. For each prevalent case, the complete process from the infection to the onset of symptoms can be considered as a renewal process. As illustrated in Figure 1, the backward recurrence time is hence defined as the time between infection and departure from Wuhan, and the forward recurrence time is the time between departure from Wuhan and symptom onset. Clearly, the forward time is observable and the corresponding observations are with good veracity, while the backward time is either unable to be observed or the corresponding observations are with large uncertainty due to recall bias. Note that for each infected individual, the backward time and forward time do not have to be same. However, when the renewal process reaches its equilibrium status, it becomes reversible, that is, the statistical properties of this process is the same as the one for time-reversed data in a same process. Hence, at equilibrium, the backward time can be treated as the forward time if time periods are reversed.^19^

**Figure 1.**
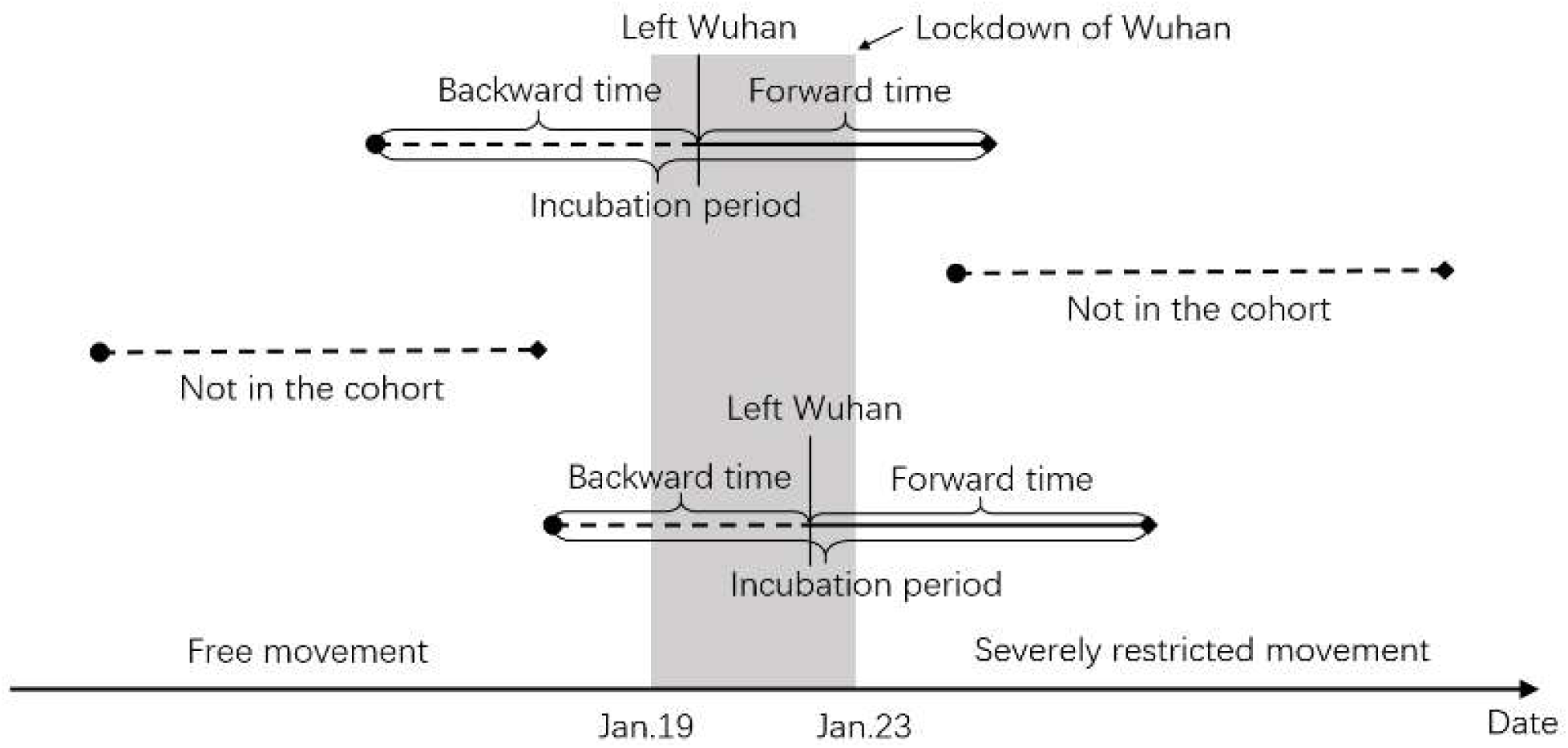
Illustration of our cross-sectional and forward follow-up study. Backward and incubation periods are not observed, while Wuhan departure and forward time are observed.

In order to model incubation using the renewal process properly, the following assumptions are established:

(A1). The renewal process has reached its equilibrium status;

(A2). The distribution of the incubation period is continuous;

(A3). The distribution of the incubation period has a finite first moment;

(A4). The incubation period for each case is independent and identically distributed;

(A5). The cases included in the analysis were infected at Wuhan and developed their symptoms outside Wuhan.

In this study, it is reasonable to assume (A1) is satisfied between January 19, 2020, and January 23, 2020, because there are over eleven million residents in the Wuhan metropolitan area and nearby neighborhoods and the daily travel volume in and out of Wuhan exceeded million before January 23, 2020. We justify the use of data between January 19 and January 23 below. With adequate long run, the renewal process would reach the equilibrium status. The assumptions (A2) to (A4) are standard. In fact, we may assume that the incubation period is a continuous variable with range (*0,M*) for some finite number *M*. It is well known that the first moment exists for a bounded random variable. The justification for assumption (A5) is below. Therefore, the probability renewal process theory can be applied with confidence, and thus we can avoid the challenging mission of ascertaining the backward time.

### Data collection and justification

Publicly available data were retrieved from provincial and municipal health commissions in China and the ministries of health in other countries, including 12 963 confirmed cases outside Hubei province as of February 15, 2020. Detailed information on confirmed cases includes region, gender, age, date of symptom onset, date of confirmation, history of travel or residency in Wuhan, and date of departure from Wuhan. The date of symptoms onset in these data refers to the date reported by the patient on which the clinical symptoms first appeared, where the clinical symptoms include fever, cough, nausea, vomiting, diarrhea, and others. Among 12 963 confirmed cases, 6345 cases had their dates of symptom onset collected; 3169 cases had histories of travel or residency in Wuhan; 2514 cases had their dates of departure recorded; and 1922 cases had records of both dates of departure from Wuhan and dates of symptoms onset.

However, not all 1922 cases should be taken in the analysis. We have to ensure that (1) assumption (A5) is satisfied, and (2) the follow-up time is long enough. To make sure that the assumption (A5) is being satisfied as much as possible, we (1) exclude cases whose first symptoms appeared before departure, and exclude cases who left Wuhan before January 19, 2020. This date was used because before January 19, the Chinese public was not aware of the severity of this epidemic, and those who left Wuhan might still have had close contact with other infected cases from Wuhan and hence actually got infected outside Wuhan. However, starting January 19, the China CDC began issuing test reagents to all provinces, confirmed cases were reported outside Hubei province in mainland China, the severity of COVID-19 was widely noted by the public, and various strict containment measures were implemented to minimize human-to-human transmission.^2^ Thus, it is unlikely that confirmed cases who left Wuhan after January 19, 2020, were infected outside Wuhan and assumption (A5) is supported. To ensure that the follow-up time is long enough such that no additional biased sampling occurred in this study, we excluded all cases who left Wuhan after January 23, 2020, which leaves an average follow-up time of 25 days (from date of departure to February 15, which is the end of this study). A 25-day follow-up period should be long enough based on the various studies on the incubation period of COVID-19.^3-6^ Note that those who left Wuhan after January 23 might not have enough time to develop symptoms before the end of the follow-up period. Including these cases in the cohort might lead to a downward bias on the incubation period. Note that the latest date of symptom onset in our cohort is February 12, 2020, which is ten days before the end of the follow-up period. This period should be long enough for a case to develop symptoms. Furthermore, there were only 49 cases who left Wuhan after the lockdown of Wuhan city on January 23, 2020.^20^ After examining the collected data, there were a total of 1211 cases that meet the criteria and were followed forwardly.

Figure 1 shows the design of the cross-sectional and forward follow-up study. The dot on the left end of each segment is a date of infection, while the square on the right end is a date of symptoms onset. Thedate of departure from Wuhan cuts the line segment in between. Note that only solid lines were followed in our cohort, while dashed lines are not in the cohort because the date of departure from Wuhan is not between January 19, 2020, and January 23, 2020.

Among the 1209 cases with gender information in the study, 533 (44·09%) are female. The mean age of patients was 41·31 and the median age was 40. Over 80% of the cases were between 20 and 60. The youngest confirmed case in our cohort was six months-old while the oldest was 86 years-old. Table 2 shows the demographic characteristics of patients with COVID-19 in the Wuhan departure cohort and the entire data collected as of February 15, 2020. We can see that there was no significant difference, which means the selected cases present the population well.

**Table 2.** Comparison between the demographic characteristics of patients with COVID-19 in the Wuhan departure cohort and all cases collected as of February 15, 2020.

| Age group (years) | Female |  | Male |  | No information |  |
| --- | --- | --- | --- | --- | --- | --- |
|  | Wuhan departure cohort | All cases | Wuhan departure cohort | All cases | Wuhan departure cohort | All cases |
|  | 533 (44.1)* | 4121 (47.3) | 676 (55.9) | 4597 (52.7) | 2 | 4245 |
| 0-20 | 17 (3.2) | 126 (3.2) | 22 (3.3) | 180 (4.2) | 0 | 3 |
| 20-39 | 215 (40.5) | 1250 (32.2) | 310 (46.3) | 1508 (35.0) | 1 | 48 |
| 40-59 | 219 (41.2) | 1667 (43.0) | 260 (38.8) | 1843 (42.8) | 0 | 57 |
| 60-79 | 76 (14.3) | 749 (19.3) | 75 (11.2) | 701 (16.3) | 0 | 40 |
| ≥80 | 4 (0.8) | 85 (2.2) | 3 (0.4) | 78 (1.8) | 0 | 8 |
| No information | 2 | 244 | 6 | 287 | 1 | 4089 |

\* Number (%). The percentages do not take missing data into account.

### Estimation of incubation period distribution of COVID-19

Let *Y* be the incubation period of an infected case with probability density function *f*(*y*) where *y*> 0. Let *V* denote the duration between departure from Wuhan and onset of symptoms, which can be considered as the forward time in a renewal process. Let *A* be the time between the infection and departure from Wuhan, which can be considered as the backward time. Clearly, *A* is not observable. It is known that in the cross-sectional sampling, *A*+*V* is a length-biased version of the incubation period *Y*, as it is easier to observe v if *A*+*V* is longer. The joint density of *A* and *V* is

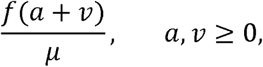

where 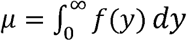 is the mean incubation period, a and v are the realizations of *A* and *V*. Marginally, *A* and *V* have the same density, that is,

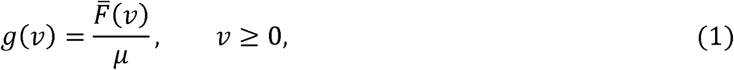

where 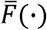 is the survival function corresponding to *f*(.) It is worth noting that *A*+ *V* is a length-biased version of *Y*, as with a longer observed *Y*, the corresponding *V* is easier to be observed. Hence the mean value *E*(*A* + *V*) is longer than the mean incubation period *µ*.

In our cohort of COVID-19 cases, we assume the incubation period is a Weibull random variable with probability density function

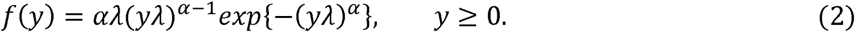

Using equations (1) and (2), it can be shown that the forward time has the density function as follows

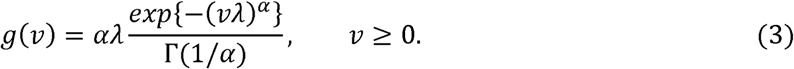

Let *v*_*i*_ be the observed forward times, *i*= 1,2,*…, I*, where *I* = 1211 in the study, the estimates 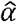 and 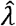 can be obtained by maximizing the likelihood function

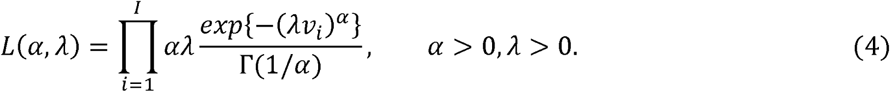

The mean and percentiles of the incubation period can be calculated from the parametric Weibull distribution. The confidence intervals in this study are obtained using bootstrap method with B = 1000 resamples.

### Sensitivity analysis

It is arguable that people who left Wuhan might also be infected at the day of departure since they had a higher chance to be exposed to this highly contagious, human-to-human–transmitted virus in a crowded environment as cases were increasing. In such case, the duration between departure from Wuhan and onset of symptoms is no longer only the forward time, but a mixture of the incubation period and the forward time. Unfortunately, it is unclear who got infected before departure and who got infected at the event of departure. Hence, a mixture sensitivity forward time model is proposed, that is,

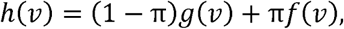

where *π* is the proportion of people who contracted disease at the event of departure. Without additional auxiliary information, in general, it is impossible to identify this model nonparametrically. Even in some fully parametric cases, *π* is not identifiable. For example, if *Y∼exp* (*λ*), that is *f*(*y*) *= λ exp* (− *y, λ*) *y > 0*, it can be shown that *V* also follows an exponential distribution with *g* (*v*) *=* (− *v λ, v* #x003E; 0. In such a case, π is not identifiable. However, in this study, with the Weibull distribution assumption, we have

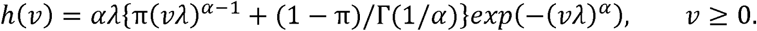

If *α* ≠ 1, it is possible to identify all underlying parameters. We explore the sensitivity of estimates of incubation period by assuming a range of π, that is π = 0, 0.05, 0.1, and 0.2, where π = 0 is the reference, and estimate *α* and *β* by maximizing the product of likelihoods, 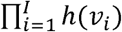, with respect to *α* and *λ*.

## Results

By fitting the observed forward times *v*_*i*_ of the 1211 cases in our cohort to the likelihood function (4), we find the maximum likelihood estimates 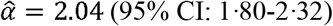 and 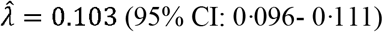 in our reference scenario. The estimated 5th, 25th, 50th, 75th, 90th, 95th, 99th, and 99·9th percentiles of the incubation period are 2·27 (95% CI: 1·73-2·86), 5·28 (95% CI: 4·53-6·06), 8·13 (95% CI: 7·37-8·91), 11·42 (95% CI: 10·74-12·11), 14·65 (95% CI: 14·00-15·26), 16·67 (95% CI: 15·94-17·32), 20·59 (95% CI: 19·47-21·62), and 25·12 (95% CI: 23·35-26·87) days, respectively. The mean incubation period is 8·62 (95% CI: 8·02-9·28) days. The average time from leaving Wuhan to symptom onset is 5·44 days, the sample median is 5 days, and the maximum is 21 days. Figure 2 visualizes the fitted density function (3) in a solid line onto the histogram of observed forward times, and the dashed line is the Weibull probability density function (2) for incubation period distribution. Note that (3) is a monotonically decreasing function, which fits the observed forward times well, suggesting that our model is reasonable and the results are therefore trustworthy.

**Figure 2.**
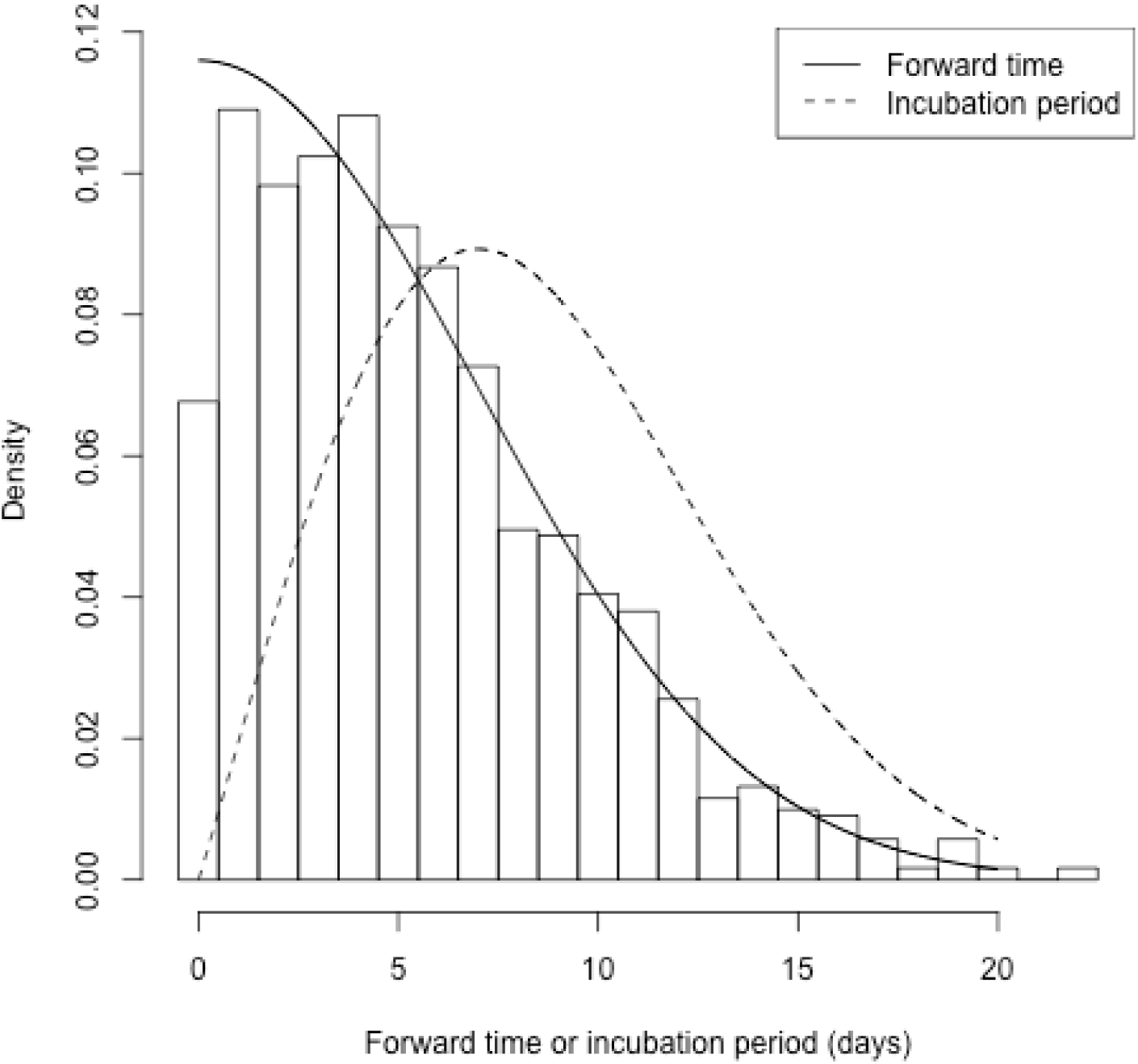
Histogram and estimated probability density functions for the time from Wuhan departure to symptoms onset, i.e., forward time.

Table 3 summaries the estimates of parameters, and the mean and percentiles of incubation period. We can see that the estimates for mean and percentiles decrease as the proportion of people who got infected at the event of departure, π, increases. However, the results are not shifted significantly. Variation of the results from π = 0 to 0.2 is only about one day.

**Table 3.** Results of our model based on different choices of.

| Scenario | Reference case | Additional % infected on the Wuhan departure day, |  |  |
| --- | --- | --- | --- | --- |
|  | 2·04 (1·80, 2·32) | 1·99 (1·78, 2·23) | 1·94 (1·76, 2·16) | 1·86 (1·71, 2·04) |
|  | 0·10 (0·10, 0·11) | 0·11 (0·10, 0·12) | 0·11 (0·10, 0·12) | 0·12 (0·11, 0·13) |
| Mean | 8·62 (8·02, 9·28) | 8·32 (7·72, 8·89) | 8·04 (7·55, 8·55) | 7·57 (7·14, 7·99) |
| 5% | 2·27 (1·73, 2·86) | 2·11 (1·67, 2·62) | 1·97 (1·60, 2·42) | 1·73 (1·42, 2·09) |
| 25% | 5·28 (4·53, 6·06) | 5·02 (4·35, 5·69) | 4·78 (4·23, 5·36) | 4·36 (3·90, 4·86) |
| Median | 8·13 (7·37, 8·91) | 7·81 (7·09, 8·50) | 7·51 (6·94, 8·11) | 7·00 (6·49, 7·50) |
| 75% | 11·42 (10·74, 12·11) | 11·06 (10·41, 11·66) | 10·73 (10·15, 11·27) | 10·16 (9·64, 10·62) |
| 90% | 14·65 (14·00, 15·26) | 14·27 (13·67, 14·85) | 13·93 (13·36, 14·45) | 13·34 (12·81, 13·87) |
| 95% | 16·67 (15·94, 17·32) | 16·28 (15·62, 16·91) | 15·94 (15·26, 16·60) | 15·37 (14·75, 16·01) |
| 99% | 20·59 (19·47, 21·62) | 20·21 (19·23, 21·26) | 19·89 (18·85, 20·87) | 19·36 (18·36, 20·37) |
| 99.9% | 25·12 (23·35, 26·87) | 24·77 (23·22, 26·43) | 24·50 (22·84, 26·11) | 24·08 (22·44, 25·71) |

## Discussion

A sound estimate of the distribution of the incubation period plays a vital role in epidemiology. Its application includes decisions regarding the length of quarantine for prevention and control, dynamic models that accurately predict the disease process, and determining the contaminated source in foodborne outbreaks. In this paper, we propose a novel method to estimate the incubation distribution which only requires information on travel histories and dates of symptoms onset. This method enhances the accuracy of estimation by reducing recall bias and utilizing abundance of the readily available forward time data. In addition, this is the first article to consider the incubation period for COVID-19 virus as a renewal process which is a well-studied methodology and has a solid theoretical foundation. The estimated incubation period has a median of 8·13 days (95% CI: 7·37-8·91), a mean of 8·62 days (95% CI: 8·02-9·28), the 90th percentile is 14·65 days (95% CI: 14·00-15·26), and the 99th percentile is 20·59 days (95% CI: 19·47-21·62). Compared with the results published in Li et al, Guan et al, Backer et al, and Linton et al, the incubation period estimated in our study is significantly longer.^3–6^ Below is some evidence that may potentially support our findings of the long incubation period:

1. In the study of Guan et al on behalf of China Medical Treatment Expert Group for COVID-19, the incubation period had a reported median of 4 days, the first quartile of 2 days and the third quartile of 7 days. ^4^ By fitting a commonly used Weibull distribution to such quartiles, we can obtain 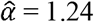 and 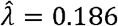 defined in Equation (2). As a consequence, the estimated 90%, 95% and 99% percentiles are, respectively, 10·54, 13·04 and 18·45 days, which indicates that some patients may have extended incubation periods.
2. One particular case reported by Yinbin municipal health commissions in China stated that a 64-year-old female was diagnosed with COVID-19 on February 11, 2020 at Yinbin, Sichuan province 20 days after returning from Wuhan. This patient was under self-quarantine at home with the family for 18 days, from January 23 to February 9. On February 8, the patient developed mild symptoms of cough with sputum production.^21^

Based on the estimated incubation distribution in this study, about 10% of patients with COVID-19 would not develop symptoms until 14 days after infection Our approach does require that certain assumptions be met, which we detail below.

1. The collection of forward time depends on the follow-up time, that is, if the follow-up time is not long enough, we would only be able to include those with a shorter incubation period in the Wuhan departure cohort. This may lead to an underestimation of the incubation period. The same limitation also applies to Backer et al and Linton et al.^5,6^ However, as explained earlier, we only included cases who left Wuhan before January 23 in this study, which leaves an average follow-up time of 25 days. Hence it is less likely we missed those patients with longer incubation periods based on the largest incubation period of 24 days reported in Guan et al.^4^ Note that the 24-day incubation period was reported as an outlier in Guan et al.^4^
2. We assume that the individuals included in our cohort were either infected in Wuhan or on the way to their destination from Wuhan, violation of such assumption lead to an overestimation of incubation period. The same limitation also applies to Backer et al and Linton et al.^5,6^ However, with a carefully selected cohort justified in the section of Method, the chance for an individual in the Wuhan departure cohort getting infected outside Wuhan should be relatively small. Nonetheless, we acknowledge this possibility exists, for example, a family member could be uninfected by the time of departing Wuhan but got infected by other family members or outside contacts after leaving Wuhan. A sensitivity analysis was also conducted by removing all cases who left Wuhan with their families in the Wuhan departure cohort, and we found it only resulted in a small change of the estimated distribution of the incubation period.
3. Individuals in our selected cohort were those who got infected in the early days of the outbreak. They were likely the first-or second-generation cases. Our results do not apply to higher generation cases if the virus mutates.

## Data Availability

data is available upon request

## Acknowledgement

We thank Dr. Dean Follmann from National Institute of Allergy and Infectious Diseases for comments that greatly improved the manuscript and Dr. Weigong Zhou from U.S. Centers for Disease Control and Prevention (CDC) for many helpful comments and suggestions. We also thank Benjamin Snow, ELS, from Leidos Biomedical Research, Inc for providing a technical review of the manuscript. This research is supported by National Natural Science Foundation of China grant 8204100362 and Zhejiang University special scientific research fund for COVID-19 prevention and control.

## Contributors

Jing Qin: study design, writing, data interpretation

Chong You: writing, literature search, data interpretation

Qiushi Lin: writing, data analysis, data collection

Taojun Hu: data analysis, data collection

Shicheng Yu: data interpretation

Xiao-Hua Zhou: study design, writing, data interpretation

## Data sharing

Data are submitted.

